# Intensive Systolic Blood Pressure Treatment Remodels Brain Perivascular Spaces: A Secondary Analysis of SPRINT

**DOI:** 10.1101/2023.02.22.23286329

**Authors:** Kyle C. Kern, Ilya M. Nasrallah, Robert Nick Bryan, David M. Reboussin, Clinton B. Wright

## Abstract

**Background:** Brain perivascular spaces (PVS) are part of the glymphatic system and facilitate clearance of metabolic byproducts. Since enlarged PVS are associated with vascular health, we tested whether intensive systolic blood pressure (SBP) treatment affects PVS structure.

**Methods:** This is a secondary analysis of the Systolic PRessure INTervention (SPRINT) Trial MRI Substudy: a randomized trial of intensive SBP treatment to goal < 120 mm Hg vs. < 140 mm Hg. Participants had increased cardiovascular risk, pre-treatment SBP 130-180, and no clinical stroke, dementia, or diabetes. Brain MRIs acquired at baseline and follow-up were used to automatically segment PVS in the supratentorial white matter and basal ganglia using a Frangi filtering method. PVS volumes were quantified as a fraction of the total tissue volume. The effects of SBP treatment group and major antihypertensive classes on PVS volume fraction were separately tested using linear mixed-effects models while covarying for MRI site, age, sex, black race, baseline SBP, history of cardiovascular disease (CVD), chronic kidney disease, and white matter hyperintensities (WMH).

**Results:** For 610 participants with sufficient quality MRI at baseline (mean age 67±8, 40% female, 32% black), greater PVS volume fraction was associated with older age, male sex, non-Black race, concurrent CVD, WMH, and brain atrophy. For 381 participants with MRI at baseline and at follow-up (median = 3.9 years), intensive treatment was associated with decreased PVS volume fraction relative to standard treatment (interaction coefficient: -0.029 [-0.055 to -0.0029] p=0.029). Reduced PVS volume fraction was also associated with exposure to calcium channel blockers (CCB) and diuretics.

**Conclusions:** Intensive SBP lowering partially reverses PVS enlargement. The effects of CCB use suggests that improved vascular compliance may be partly responsible. Improved vascular health may facilitate glymphatic clearance.

Clincaltrials.gov: NCT01206062

## Introduction

Brain perivascular spaces (PVS) are visible on routine MRI and are an emerging biomarker for cerebral small vessel disease (CSVD). Enlargement of PVS is thought to reflect stagnant flow, and is associated with aging, hypertension, and other cerebrovascular risk factors.^1-3^ PVS are part of the glymphatic system, which facilitates the exchange and clearance of solutes between the cerebrospinal fluid and interstitial fluid.^4^ Based on preclinical models, fluid movement in the PVS is thought to be partly driven by arterial pulsations and low frequency vasomotor oscillations.^4-7^ Thus, arterial stiffening associated with arteriolosclerosis and aging potentially impairs bulk flow, leading to PVS enlargement and impaired glymphatic clearance.^6, 7^ Since glymphatic circulation facilitates clearance of protein aggregates including amyloid,^8^ enlarged PVS may reflect a process by which poor vascular health contributes to neurodegenerative disease.^4^

Most prior studies have relied on visual rating scales to quantify PVS,^9^ but volumetric quantification may be more sensitive to longitudinal changes. Established methods permit unbiased calculation of PVS volumes^10, 11^ that have shed light on the dynamic nature of PVS in development,^12^ cognitive impairment,^13^ Parkinson’s disease,^14^ and space flight.^15^

While enlarged PVS are associated with hypertension^2^, whether antihypertensive therapy and intensive blood pressure treatment can affect PVS structure is unknown. The SPRINT trial evaluated the effects of intensive systolic blood pressure control, targeting SBP less than 120 mm Hg vs. the standard therapy goal of less than 140 mm Hg, on cardiovascular and cognitive outcomes and, in a subsample, brain imaging markers. In this subsample, intensive therapy resulted in slower progression of the CSVD biomarker of WMH volume.^16, 17^ In this secondary analysis of SPRINT we evaluate the effect of intensive treatment on PVS volumes. We hypothesized that automated PVS segmentation could detect longitudinal changes in PVS volume, and that intensive treatment would slow the enlargement of PVS.

## Methods

### Data Availability

Anonymized data used in this study are available in the NHLBI data repository BioLINCC (biolincc.nhlbi.nih.gov). Additional data that support the findings of this study are available from the corresponding author upon reasonable request.

### Trial Design

The trial design, methods, primary outcomes, and protocol have been published previously,^16-18^. The trial and MRI substudy were approved by the institutional review board at each participating site, and each participant provided written informed consent.

### Participants

Participants were recruited from clinic settings or from the community. Participants were ≥ 50 years old with SBP between 130 and 180 mm Hg at screening and had increased cardiovascular risk. Increased cardiovascular risks included clinical or subclinical cardiovascular disease (CVD), chronic kidney disease (CKD), a Framingham risk score of ≥ 15%, or age ≥ 75 years. Participants were excluded if they had diabetes, a history of stroke, a diagnosis of dementia or were treated with dementia medications, or lived in a nursing home. Race and ethnicity were collected via self-report as recommended by National Institutes of Health Guidelines. Participants were randomized by the data coordinating center, stratified by clinic site, in a 1:1 ratio to either an SBP goal < 120 mm Hg (intensive treatment; n = 4678) or an SBP goal < 140 mm Hg (standard treatment; n = 4683). The algorithms and formulary for antihypertensive treatment have previously been published,^18^ and included all major classes, provided at no cost to participants. The protocol encouraged but did not mandate thiazide diuretics as first-line, loop diuretics for participants with CKD, and Beta-adrenergic blockers for participants with coronary artery disease.

### Antihypertensive classes

Antihypertensive medications were recorded at each study visit and exposure to each class was calculated as the fraction of days taking the antihypertensive from randomization to follow-up MRI. To test the effects of antihypertensive classes on PVS volumes, some classes were combined: ACE inhibitors and angiotensin receptor blockers, selective and non-selective beta blockers, dihydropyridine and non-dihydropyridine calcium channel blockers, and diuretics including thiazide, loop, and potassium-sparing.

### Achieved Systolic Blood Pressure

All blood pressure measures from randomization to the follow-up MRI were used to calculate the achieved SBP, which was calculated as the area under the SBP curve divided by the number of days.

### MRI Acquisition

Exclusion criteria for the MRI substudy included contraindications to MRI such as implanted or foreign metallic or ferromagnetic material, or severe claustrophobia. Multimodal brain MRI was obtained at baseline and planned at 48 months post-randomization, both performed on the same scanner. Due to early termination of the study due to improved cardiovascular outcomes with intensive treatment, follow up MRIs were obtained earlier than planned. MRIs were obtained at 7 sites using 3T scanners (3 Phillips and 4 Siemens). The protocol included 1mm isotropic 3D T1, T2 and FLAIR sequences. Enrollment continued from November 8, 2010 to March 2013, with the last MRI occurring in July 2016. Of 1267 individuals screened, 793 were eligible, and 662 had baseline T1 and T2 that passed initial quality control.

### MRI Processing

Image parameters were calculated as previously described, including white matter hyperintensity volume (WMH), total brain volume (TBV), total white matter (WM) volume, total gray matter (GM) volume, and total intracranial volume (TICV).^17^ WMH volumes were adjusted for TICV and log transformed due to skewed distribution (logWMH = log(1 + WMHvol/TICV). Brain parenchymal fraction (BPF), a marker of atrophy, was calculated as BPF= (WM + GM) / TICV.

### Calculation of Perivascular Space Volumes

PVS volumes were calculated using T1, T2 and FLAIR MRI following methods established by Sepehrband et al.^11^ and Ballerini, et al.^10, 19^ using FreeSurfer (surfer.nmr.mgh.harvard.edu/fswiki/Samseg),^20^ FSL (fsl.fmrib.ox.ac.uk/fsl),^21^ and the Quantitative Imaging Toolkit (cabeen.io/qitwiki).^22^

T1, T2 and FLAIR images were bias-field corrected and aligned, and tissue was segmented using a multi-modality tissue segmentation algorithm to create a region-of-interest (ROI) of the supratentorial white matter and basal ganglia^20^ for PVS segmentation. The ROI mask was smoothed with a 1mm filter while retaining edges, and a hole-filling algorithm was applied to fill regions where PVS were labelled as CSF. A ventricular mask was dilated by 1mm to further remove the layer of voxels immediately adjacent to the ventricles from this ROI.^11^

T2 images were used to segment PVS within this ROI (Figure 1). First a non-local means filter was applied to reduce Rician noise. Next a Frangi filter was applied that selects for hyperintense, tubular structures and assigns a value of “vesselness” using parameters previously established:^11^ alpha=0.5, beta=0.5, gamma=500, Smin=0.1, and Smax=0.5. Next, the Frangi intensity map was scaled to the interquartile range to avoid the influence of large outliers and then thresholded using a value of 2.7, which was determined from the dataset as described below. WMH were excluded from the PVS segmentation. Finally, the PVS volume is calculated in cm^3^ and also reported as a percentage of the total ROI volume analyzed (white matter and basal ganglia): the PVS volume fraction.

**Figure 1.**
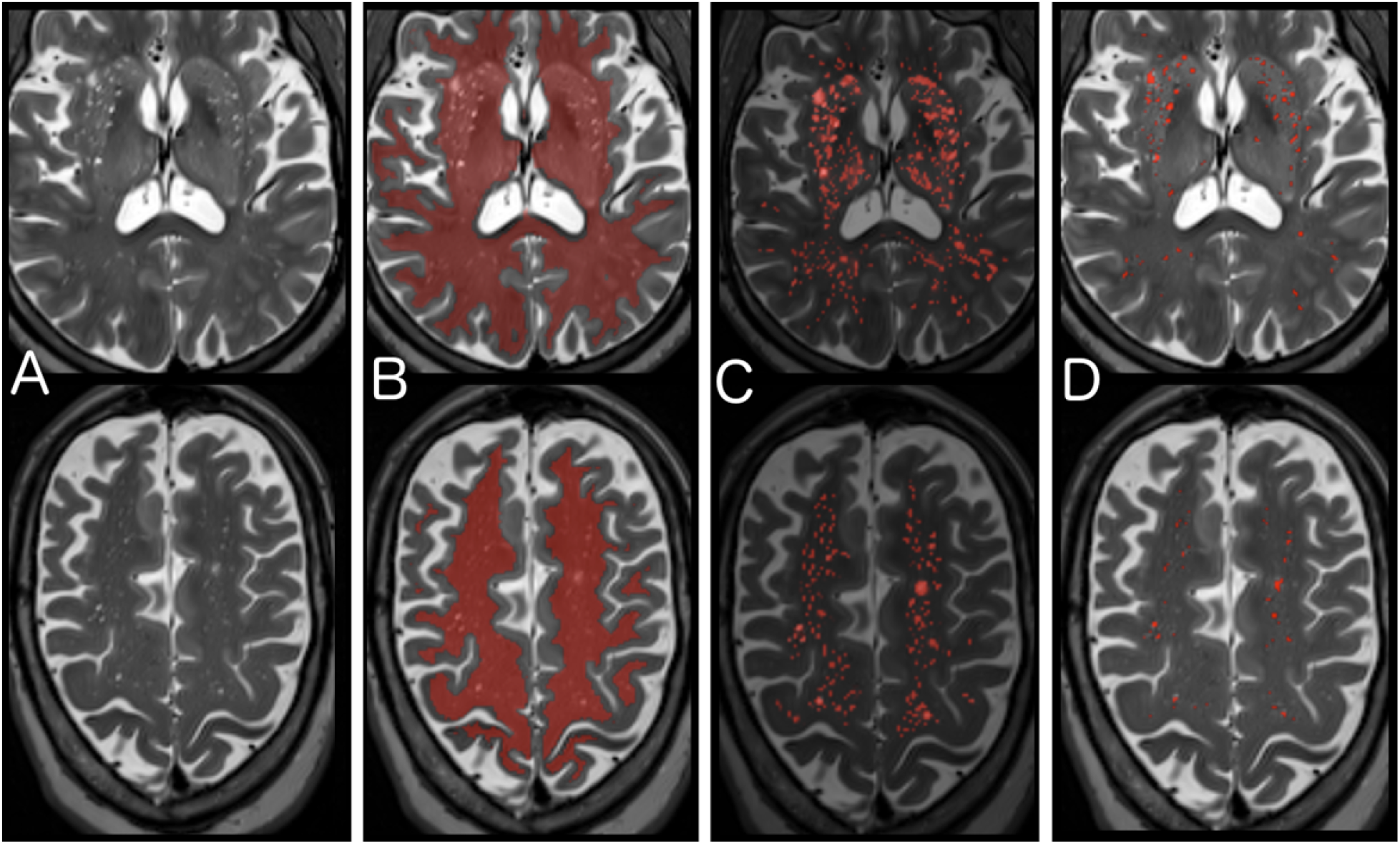
Segmentation of Perivascular Spaces. A) T2 images are corrected for field inhomogeneity, and then a non-local means filter is applied that reduces Rician noise. B) A region of interest (ROI) of supratentorial white matter and basal ganglia is created from the T1 and FLAIR images using Freesurfer ‘s SAMSEG^20^ and aligned to the T2 image. C) A Frangi filter is applied within the ROI that selects for tubular structures and assigns an intensity reflecting the “vesselness” at each voxel. D) The optimal Frangi threshold of 2.7 (standardized value) was determined empirically by comparing to visual ratings and visually confirmed for anatomical match across a random subset of 120 images.

**Figure 2.**
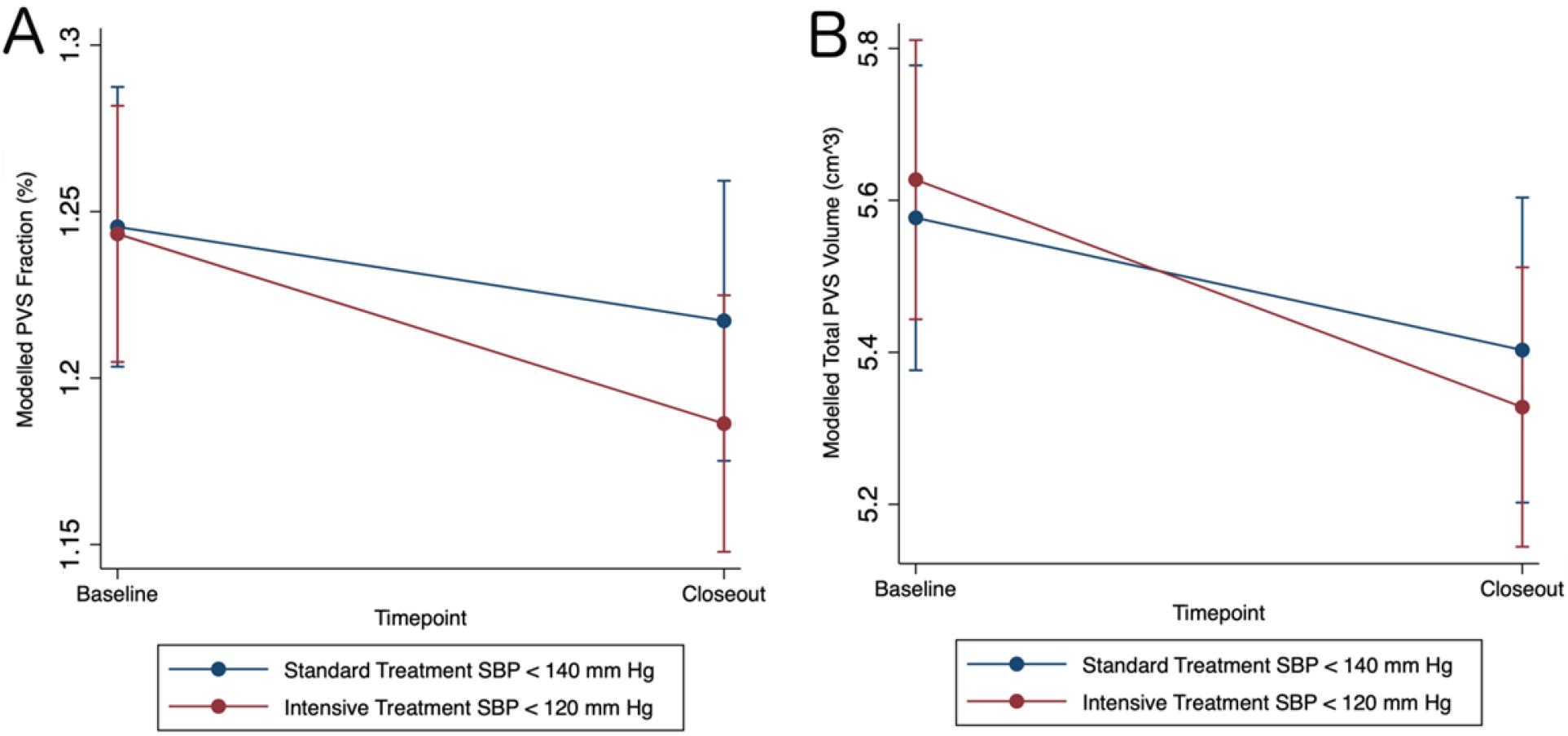
Intensive Treatment Reduces PVS Volumes. A) Modelled perivascular space (PVS) volume fraction from over time by systolic blood pressure (SBP) treatment group. B) Modelled total PVS volume over time by SBP treatment group.

To determine the optimal scaled Frangi threshold for T2 images in this dataset, a random sample of 120 scans were used to compare visual PVS ratings to calculated PVS volumes across a range of Frangi thresholds. Supratentorial PVS were visually rated on T2 using an established grading system^9^ for this sample by one rater and then repeated after a 2-month washout period. PVS were rated in the basal ganglia and insular region (0 to 4), and the centrum semiovale (0 to 4), providing a total supratentorial visual PVS score of 0 to 8. For these 120 scans, the Frangi image threshold was varied from 0.5 to 3, and a correlation coefficient was calculated at each threshold between the calculated percent volume fraction and the visual rating score. The optimal threshold of 2.7 (R=0.510) was determined by considering the maximal correlation coefficient and visual inspection (supplemental figure 2).^11^ Finally, PVS were segmented for all image sets in a fully automated fashion using a high-performance computing cluster (hpc.nih.gov), and then visually inspected (while blinded to treatment and timepoint) to ensure quality segmentation. Participants with T2 images that were of insufficient quality to visualize and segment PVS were excluded (N= 105: 52 at baseline and 53 at follow-up).

### Covariates and Subgroups

Prespecified covariates for the trial’s primary outcomes included age, sex, race (Black vs not Black), CKD subgroup (eGFR < 60 vs ≥ 60 mL/min/1.73 m^2^), history of CVD, and baseline SBP. In the baseline analysis, the relationship to BPF, a marker of atrophy was tested. Since severe WMH can obscure PVS and their segmentation, logWMH was also included as a covariate.

### Statistical Analysis

Predictors of baseline PVS volume fraction were determined using multiple linear regression. Mixed effects linear regression was used to estimate change in PVS volume fraction over time between treatment groups. Random effects included participant and MRI facility, while fixed effects included timepoint, treatment group, age, sex, race, CVD subgroup, CKD subgroup, baseline SBP, and logWMH. The primary outcome was the interaction effect between treatment group and timepoint, where the coefficient reflects the relative change in PVS volume fraction between treatment groups (Model 1). Secondarily, pairwise comparisons were tested, and Bonferroni corrected, to determine absolute differences between each timepoint and group. A similar mixed effects model was used to test whether change in total PVS volume was related to treatment (Model 2). Fixed effects included timepoint, treatment, TICV, WMH volume, baseline SBP, age, sex, race, CVD and CKD. The interaction between time and treatment group reflects the relative change in PVS volumes between treatment groups. Again, pairwise comparisons, Bonferroni corrected, were used to test absolute differences over time and between groups. Finally, the effects of antihypertensive classes on PVS were tested with a model that also included fixed effects for exposure to each of 4 major antihypertensive classes: ACE inhibitors or ARBs (ACEi/ARB), beta-blockers, calcium channel blockers (CCB), or diuretics. Additional covariates included treatment group, baseline SBP, achieved SBP, age, sex, race, CVD, CKD and logWMH. Four interaction terms between timepoint and antihypertensive class exposure were included to test the effect of each class on change in PVS volume fraction (Model 3). We also reported the baseline differences between those with and without follow-up MRI (supplemental table 1). All hypothesis tests were 2-sided, and adjusted p-values less than 0.05 were considered statistically significant. Analyses were performed March 2022 through February 2023 using Stata v.17.0 (stata.com).

## Results

To determine baseline predictors of PVS volume fraction, 610 participants were included (table 1). For the longitudinal analysis, 381 participants had both baseline and follow-up MRI of sufficient quality: 207 with intensive treatment and 174 with standard treatment (Supplemental Figure 1).

**Table 1.** Baseline Clinical and Imaging Measures

|  | Overall | Intensive Tx | Standard Tx |
| --- | --- | --- | --- |
| N | 610 | 324 | 286 |
| % Female | 40% (245) | 44% (141) | 36% (104) |
| Age (years ± SD) | 68 ± 8 | 68 ± 8 | 67 ± 9 |
| <u>Race:</u> |  |  |  |
| Black | 32% (198) | 31% (102) | 34% (96) |
| White | 66% (403) | 67% (217) | 65% (186) |
| Hispanic | 5% (30) | 3% (11) | 7% (19) |
| Other | 2% (12) | 2% (6) | 2% (6) |
| Baseline SBP (mm Hg) | 138 ± 16 | 137 ± 17 | 137 ± 16 |
| Framingham Score (± SD) | 17 ± 2 | 17 ± 2 | 17 ± 2 |
| <u>Self-Reported History of:</u> |  |  |  |
| Hypertension | 93% (569) | 94% (305) | 92% (264) |
| Smoked > 100 lifetime cigs | 53% (324) | 54% (175) | 52% (149) |
| Taking Aspirin | 51% (312) | 49% (158) | 54% (154) |
| Cancer | 13% (80) | 15% (48) | 11% (32) |
| Current Smoking | 12% (77) | 13% (43) | 12% (34) |
| Atrial Fibrillation | 7% (40) | 7% (23) | 6% (17) |
| Peripheral Vascular Disease | 6% (34) | 7% (23) | 4% (11) |
| Heart Attack | 4% (27) | 5% (15) | 4% (12) |
| Prior TIA | 3% (20) | 4% (13) | 2% (7) |
| Congestive Heart Failure | 2% (14) | 2% (7) | 2% (7) |
| Seizure | 1% (7) | 1%(3) | 1% (4) |
| <u>Baseline Imaging Measures:</u> |  |  |  |
| WMH Volume (cm<sup>3</sup> ± IQR) | 3.2 ± 4.6 | 3.1 ± 5.1 | 3.3 ± 4.4 |
| Brain Parenchymal Fraction (%) | 82 ± 4 | 82 ± 4 | 82 ± 4 |
| PVS Volume (cm<sup>3</sup> ± SD) | 5.49 ± 1.70 | 5.45 ± 1.74 | 5.52 ± 1.67 |
| PVS Volume Fraction (% ± SD) | 1.23 ± 0.32 | 1.23 ± 0.33 | 1.23 ± 0.31 |

WMH: white matter hyperintensity. PVS: perivascular space. SD: standard deviation

PVS visual ratings were performed for 120 randomly selected participants to determine the optimal Frangi threshold and repeated twice by the same rater separated by 2 months. Intraclass correlation coefficient for intra-rater PVS visual rating was 0.80. PVS volume fractions were associated with PVS visual ratings for the centrum semiovale and basal ganglia combined (R= 0.51) (Supplemental Figure 2). Quality review was also repeated for this sample resulting in a Kappa of 1.0 for scan exclusion/inclusion.

Baseline characteristics were similar between the intensive and standard SBP treatment groups (Table 1). Larger PVS volume fraction at baseline was associated with older age, male sex, non-black race, CVD subgroup, lower logWMH, larger BPF while controlling for MRI site (Table 2). Over median 3.9 years follow-up, mean achieved SBP was 120 ± 8 mm Hg with intensive treatment and 136 ± 7 mm Hg with standard treatment (p<0.001) (Supplemental Figure 3). At baseline, PVS volume fractions were 1.23% in both groups while total PVS volumes were 5.45 cm^3^ in the intensive treatment group and 5.52 cm^3^ in the standard treatment group (Table 1).

**Table 2.** Baseline Predictors of PVS Volume Fraction.

| **Predictors:** | **Coefficient** | **95% C.I.** | **Std. Beta** | **T-statistic** | **p-value** |
| --- | --- | --- | --- | --- | --- |
| Male Sex | 0.081 | 0.039 to 0.12 | 0.12 | 3.81 | **<0.001** |
| BPF | 0.012 | 0.0061 to 0.020 | 0.15 | 3.69 | **<0.001** |
| Age | 0.004 | 0.0085 to 0.0072 | 0.10 | 2.50 | **0.013** |
| CVD Subgroup | 0.067 | 0.0097 to 0.012 | 0.072 | 2.30 | **0.022** |
| Baseline SBP | 0.0025 | -0.00092 to 0.0014 | 0.013 | 0.42 | 0.67 |
| CKD Subgroup | -0.021 | -0.065 to 0.023 | -0.029 | -0.93 | 0.35 |
| Black Race | -0.065 | -0.11 to -0.021 | -0.095 | -2.89 | **0.004** |
| logWMH | -1.05 | -1.22 to -0.89 | -0.42 | -12.84 | **<0.001** |

Adjusted for MRI site. BPF = Brain Parenchymal fraction. CVD = Cardiovascular Disease. SBP = Systolic Blood Pressure. CKD = Chronic Kidney Disease. WMH = White Matter Hyperintensities.

In the longitudinal analysis, there was a reduction in mean PVS volume fraction in the intensive treatment group relative to the standard treatment group over time by -0.029% (−0.055 to -0.0029, p=0.029) (Table 4, Model 1). In pairwise comparisons testing absolute group and timepoint differences, the intensive group had a -0.027% decrease in PVS volume fraction from baseline (−0.51 to -0.0032, p=0.016) while the standard group had a non-significant decrease of -0.0017% (−0.025 to 0.029).

We next tested whether treatment was associated with change in the total PVS volume while covarying for the total WMH volume and total intracranial volume. Relative to standard treatment, intensive treatment was associated with -0.13 cm^3^ decrease in PVS volume (−0.25 to -0.0026, Table 4, Model 2). The absolute change from baseline with intensive treatment was -0.20 cm^3^ (−0.31 to -0.089, p < 0.001) and -0.076 cm^3^ with standard treatment (−0.20 to 0.050).

Finally, we tested the effects of the 4 major antihypertensive classes on mean PVS volume fraction. ACEi/ARBs were the most common class of antihypertensive at baseline, and participants were also most exposed to this class during the study. CCBs were the most frequently added medication during the study (Table 3). At baseline there were no associations between antihypertensive use and PVS volume fractions. In the longitudinal analysis, longer exposure to CCBs and diuretics were both associated with greater reduction in PVS volume fraction while also covarying for the other major classes, treatment group, and achieved SBP. While maximum CCB exposure (e.g. 100% study duration) was associated with -0.036% reduction in PVS volume fraction (−0.066 to -0.0059, p=0.019), maximum diuretic exposure was associated with -0.031% PVS volume fraction reduction (−0.061 to -0.0012, p=0.041) (Table 4, Model 3).

**Table 3.** Antihypertensive Use

|  | Baseline | Longitudinal Analysis |  |
| --- | --- | --- | --- |
|  |  | Newly Added | Exposure ≥ 1 Year |
| **n** | 610 | 381 | 381 |
| **ACEi or ARB** | 70% (424) | 29% (112) | 80% (307) |
| **ACE inhibitor** | 45% (277) | 14% (55) | 44% (170) |
| **ARB** | 28% (168) | 20% (78) | 41% (158) |
| **Diuretics** | 61% (371) | 19% (73) | 68% (265) |
| **Thiazide Diuretic** | 56% (340) | 18% (70) | 62% (237) |
| **K+ Sparing Diuretic** | 10% (63) | 13% (49) | 16% (63) |
| **Loop Diuretic** | 4% (22) | 5% (21) | 7% (26) |
| **Calcium Channel Blocker** | 40% (246) | 35% (136) | 58% (224) |
| **Dihydropyridine** | 34% (208) | 34% (130) | 53% (204) |
| **Non-Dihydropyridine** | 6% (39) | 3% (13) | 6% (24) |
| **Beta Blocker** | 35% (215) | 12% (48) | 37% (143) |
| **Alpha-1 Blocker** | 3% (20) | 8% (32) | 6% (22) |
| **Alpha-2 Agonist** | 2% (12) | 3% (10) | 2% (8) |
| **Hydralazine** | 1% (6) | 8% (30) | 4% (15) |
| **Nitrate** | <1% (1) | <1% (2) | <1% (3) |
| **<u>Number of Anithypertensive Classes</u>** |  |  |  |
| **0** | 2% (11) | 30% (116) | 2% (6) |
| **1** | 22% (137) | 34% (134) | 14% (54) |
| **2** | 38% (232) | 23% (90) | 29% (111) |
| **3** | 28% (175) | 9% (34) | 30% (115) |
| **4** | 8% (51) | 2% (9) | 19% (73) |
| **5** | < 1 % (4) | <1% (2) | 6% (22) |
| **6+** | 0 | 0 | 1% (4) |

**Table 4.** Regression Models for Longtiudinal Perivascular Space Changes

| **Model 1: Effect of Treatment on PVS Volume Fraction (n=381)** |  |  |  |
| --- | --- | --- | --- |
| <u>Fixed Effects Predictors:</u> | coefficient | 95% CI | p-value |
| Timepoint | -0.0017 | -0.018 to 0.022 | 0.87 |
| Intensive Treatment | -0.0021 | -0.059 to 0.055 | 0.94 |
| <u>Interaction:</u> |  |  |  |
| Intensive Treatment x Time | -0.029 | -0.055 to -0.0029 | **0.029** |
| Adjusted for: age, sex, race, CVD, CKD, baseline SBP, MRI site, and logWMH |  |  |  |
| **Model 2: Effect of Treatment on PVS Volume (n=381)** |  |  |  |
| <u>Fixed Effects Predictors:</u> | coefficient | 95% CI | p-value |
| Timepoint | -0.076 | -0.17 to 0.018 | 0.113 |
| Intensive Treatment | 0.05 | -0.22 to 0.32 | 0.72 |
| <u>Interactions:</u> |  |  |  |
| Intensive Treatment x Time | -0.13 | -0.25 to -0.0026 | **0.045** |
| Adjusted for: age, sex, race, CVD, CKD, baseline SBP, MRI site, WMH volume, and total intracranial volume |  |  |  |
| **Model 3: Effect of Antihypertensive Class (n=381)** |  |  |  |
| <u>Fixed Effects Predictors:</u> | coefficient | 95% CI | p-value |
| Timepoint | 0.02 | -0.060 to 0.017 | 0.27 |
| <u>Interactions:</u> |  |  |  |
| ACEi / ARB x Time | -0.0022 | -0.036 to 0.031 | 0.90 |
| Beta Blockers x Time | -0.0086 | -0.021 to 0.038 | 0.57 |
| CCBs x Time | -0.036 | -0.066 to -0.0059 | **0.019** |
| Diuretics x Time | -0.031 | -0.061 to -0.0012 | **0.041** |
| Adjusted for: age, sex, race, CVD, CKD, baseline SBP, treatment group, Δachieved SBP, MRI site, logWMH, and main effects for exposures to ACEi/ARB, Beta blockers, CCB, and diuretics | | | |

PVS: perivascular spaces. CI: confidence intervals. CVD: cardiovascular disease. CKD: chronic kidney disease. SBP: systolic blood pressure. WMH: white matter hyperintensities. ACEi: ace inhibitors. ARB: angiotensin receptor blockers. CCBs: calcium channel blockers.

## Discussion

Intensive treatment to goal SBP < 120 mm Hg reduced PVS volume fraction by 0.029 percentage points relative to standard treatment, with an absolute reduction of -0.027 percentage points (−0.20 cm^3^) from baseline. When compared to the baseline association with age (0.004 percentage points larger per year older), the reduction in PVS volume fraction from baseline equated to an approximately 7-year reversal in age-related PVS enlargement (over median 3.9 years). Interestingly, even the standard treatment group showed a nonsignificant reduction of -0.0017 percentage points (−0.075 cm^3^) from baseline, which was still an attenuation of the effect of age on baseline PVS volume fraction. The reduction in PVS volume fraction was also associated with exposure to both CCBs and diuretics, but not the use of other antihypertensive classes, suggesting potential mechanisms for PVS changes. The implications of these findings are that intensive treatment alters PVS morphology, partially reversing the PVS enlargement seen with aging, hypertension, and increased vascular risk. It remains to be seen whether these SBP-driven PVS alterations reflect improved glymphatic function and brain health or are merely an epiphenomenon of intensive SBP treatment.

The primary result of the SPRINT MRI substudy was that intensive treatment slowed progression of WMH.^17^ Our results demonstrated a partial reversal of PVS enlargement, suggesting that PVS may be more dynamic than other markers of CSVD. While some reversal of WMH has been shown,^23, 24^ WMH are thought to reflect white matter injury through multiple proposed mechanisms that are largely irreversible. The primary SPRINT MRI publication also reported that intensive SBP treatment reduced total brain volume by 30.6 cm^3^ (−2.7%) compared to 26.9 cm^3^ (−2.4%) in the standard treatment group. PVS volumes did not account for this magnitude of change, being only 5.3 cm^3^ on average, and longitudinal PVS changes were much smaller (−0.20 cm^3^ vs -0.076cm^3^). On the other hand, visible PVS may reflect only a small fraction of subvoxel-sized PVS and other extracellular fluid spaces regulated via the glymphatic system. Conversely, total brain volume changes did not appear to completely account for the PVS change, since we used PVS volume fraction, a PVS density measure that accounted for changes to the underlying tissue.

While we detected reversal of PVS enlargement seen with aging and CSVD, there is little evidence from human studies that PVS size directly relates to glymphatic function. One potential explanation for our findings could be that overall volume status was reduced in the intensive SBP treatment group, and that fluid shifts preferentially affected PVS. In support of this, we found that diuretic exposure was associated with PVS volume reduction. However, we also found a stronger association between CCB exposure and PVS volume reduction. CCBs, particularly dihydropyridines, exert antihypertensive effects primarily through arterial vasodilation, thereby increasing vascular compliance of the cerebral arteries. Improved cerebrovascular compliance may have reduced PVS size by facilitating bulk flow in the PVS and interstitial fluid through greater amplitude vasomotor oscillations. Prior SPRINT results in a different subsample found that the progression of aortic stiffness was attenuated by intensive SBP treatment,^25^ so it is possible that intensive treatment also affected cerebrovascular stiffness. However, there were multiple potential confounders in interpreting the relative effects of antihypertensive classes, since medication exposure was not randomized. It is difficult to disentangle medication class effects from participants’ comorbidities and prescriber preferences that would bias antihypertensive exposure. For example, while thiazide diuretics were the encouraged first-line agent, CCBs were the most common new antihypertensive added during the study. Furthermore, ACEis or ARBs were used by 80% of participants, reducing the ability to detect a class effect.

The baseline associations between enlarged PVS and older age have been previously reported in studies using both visual PVS rating scales^2, 26^ and PVS quantification^27^. Some have suggested that enlarged PVS merely reflect local atrophy.^28, 29^ However, in this study, we find that enlarged PVS volume fraction was positively associated brain parenchymal fraction, or less atrophy. Although this finding is counterintuitive, the positive association suggests that PVS enlargement reflects more than just brain atrophy. While we quantified PVS as a volume fraction reflecting PVS density within the tissue studied, prior studies also found increased PVS visibility was associated to larger intracranial volume.^29^

Larger PVS volume fraction was also associated with a lower burden of WMH. This would seem to contradict multiple prior studies demonstrating increased PVS visibility in the setting of SVD and higher WMH burden.^1, 9^ However, in this volumetric assessment, the PVS segmentation method restricted any overlap with WMH. As a result, a larger burden of WMH resulted in less white matter space to quantify PVS. In this older population with elevated cardiovascular risk, more severe WMH burden was common. While PVS run through WMH, and WMH are sometimes seen to develop around PVS, the overlap is difficult to distinguish on T2, so voxels containing WMH were excluded from PVS segmentations. To adjust for this discrepancy, we included logWMH as a covariate throughout all analyses. However, since WMH growth was attenuated in the intensive treatment group in the initial SPRINT MRI study, the inverse relationship between WMH and PVS volume fraction might have reduced the effect size of intensive treatment on PVS volume reduction. Given that the physiologic relationship between enlarging PVS and WMH remains unclear, a voxel-wise, longitudinal analysis co-localizing PVS and new WMH growth could provide further insight on the role of visible PVS in CSVD progression.

There are several limitations to this study. Of participants with baseline MRI, 31% did not have follow-up MRI, thus introducing potential retention bias. While the attrition was partially affected by early termination of the intervention,^17^ loss to follow-up may have selected for healthier and more compliant participants. Furthermore, due to the small size of PVS, quantification is limited by motion and T2 image quality, and 9% of scans were excluded due to insufficient image quality. This loss was compounded for the longitudinal analysis, with 16% of participants excluded due to insufficient baseline or follow-up scans. While all images and segmentations were reviewed for quality and visually poor segmentations were excluded, there may have been a persistent effect of image quality on the volumetric segmentation of PVS in the remaining datasets. For the automatic PVS segmentation, a random subset of 120 scans were visually rated to determine the optimal threshold for the dataset, without individualizing for differences in MRI sites or local populations. While we adjusted for MRI site in all models, individualized thresholds could provide closer anatomical segmentation of PVS. However, MRI site sequences were coordinated using shared phantoms, and each participant had both scans performed at the same site. Furthermore, in determining the optimal Frangi threshold, the relationship between PVS volume fraction and visual ratings was stable over a wide range of thresholds, suggesting that small differences in thresholding would make little difference to the results.

## Conclusions

Enlarged perivascular spaces are a dynamic marker of CSVD, and intensive treatment partially reverses the PVS enlargement seen with aging and vascular risk factors. While SBP lowering contributes to PVS remodeling, we also found additional effects of exposure to calcium channel blockers and diuretics, suggesting that both improved vascular compliance and fluid shifts may contribute to reduction of PVS volumes. While it remains to be seen whether these structural changes reflect improved glymphatic function, our findings implicate a potential mechanism by which intensive SBP treatment may improve brain health.

## Data Availability

Data used in this analysis is available via request through NHLBI BioLINCC Data Repository. Additional derived quantitative data may be shared upon request to the corresponding author.

biolincc.nhlbi.nih.gov

## Acknowledgements

This work utilized the computational resources of the NIH HPC Biowulf cluster: hpc.nih.gov.

## Disclosures

KCK: None. IMN: None. NMP: None. DMR: None. CBW: Honoraria from uptodate.com for articles on vascular dementia. This article was not reviewed by the SPRINT Publications and Presentations Committee.

Supplemental Material

Table S1: Excluded Participants

Table S2: Imaging Metrics

Figure S1: Inclusion/Exclusion Flowchart

Figure S2: Determination of the Frangi Threshold

Figure S3: Achieved Systolic Blood Pressure

